# ClinGen Glaucoma Variant Curation Expert Panel recommendations enhance classification of myocilin variants

**DOI:** 10.64898/2026.07.29.26359282

**Authors:** Stuart W. J. Tompson, Patricia Graham, Johanna Hadler, Francesca Pasutto, Kristina N. Whisenhunt, Subhabrata Chakrabarti, Terri L. Young, Jamie E Craig, Alex W Hewitt, Owen M Siggs, John D Hulleman, David A. Mackey, Kathryn P. Burdon, Andrew Dubowsky, Emmanuelle Souzeau

**Author notes:** **Correspondence** Emmanuelle Souzeau, Department of Ophthalmology, Flinders University, 1 Flinders Drive, Bedford Park, SA, 5042, Australia.

## Abstract

Pathogenic variants in the *myocilin* (*MYOC*) gene are the most common cause of Mendelian open-angle glaucoma. In 2022, the Clinical Genome Resource (ClinGen) Glaucoma Variant Curation Expert Panel (VCEP) published rule specifications for *MYOC* variant interpretation, including a pilot study of 81 variants. Here, we present the results of curating 271 *MYOC* variants reported in people with open-angle glaucoma using updated specification rules.

Of all the variants, 11 were classified as benign (B), 45 as likely benign (LB), 166 as variants of uncertain significance (VUS), 35 as likely pathogenic (LP), and 14 as pathogenic (P). All LP/P variants were located within the conserved olfactomedin domain encoded by exon 3.

The updated variant curation guidelines from the Glaucoma VCEP increased the number of clinically definitive classifications from 28% (74/265) to 39% (105/271), with 95% (41/43) of reclassified variants moving to greater clinical relevance. Functional evidence was lacking for 93% (154/166) of VUS. Additional functional evidence could further enhance classification by halving (85/166) the proportion of those classified as VUS. These findings highlight the role of rule calibration and rigorous functional evidence assessment toward improving variant classification with clinical utility for patients.

## INTRODUCTION

Glaucoma is a term used to describe a diverse group of ocular diseases characterized by optic neurodegeneration and visual field loss, and represents the leading cause of irreversible blindness worldwide [1, 2]. The most common form is primary open-angle glaucoma (POAG), which is defined by an open anterior chamber angle and is often associated with elevated intraocular pressure (IOP) in adults 40 years of age or older. It typically presents as a chronically progressive disease [3, 4]. A subtype of POAG, known as juvenile open angle glaucoma (JOAG), is diagnosed before the age of 40 [5]. Glaucoma that manifests before four years of age is further delineated as primary congenital glaucoma. This form, while part of the open-angle glaucoma spectrum, usually results from abnormal development of the aqueous humor outflow pathway tissues rather than a progressive decline in their function [6, 7].

Pathogenic variants in the *myocilin* gene (*MYOC*, MIM 601652; previously known as *trabecular meshwork-induced glucocorticoid response* or *TIGR*) were the first identified molecular cause of glaucoma and are the most common Mendelian basis for JOAG and, to a lesser extent, POAG [8, 9, 10]. *MYOC*-related glaucoma follows an autosomal dominant inheritance pattern, with pathogenic variants increasing an individual’s estimated lifetime risk for developing glaucoma by 48% to 100% [11, 12, 13]. Myocilin is highly expressed in the trabecular meshwork (TM) of the anterior draining angle of the eye, which is crucial for the regulation of aqueous humor outflow. While the Myocilin protein is normally secreted into the extracellular matrix, variants associated with glaucoma induce abnormal protein folding, which lead to intracellular retention and aggregate formation [14, 15]. Histological analysis of a human donor eye with *MYOC*-related glaucoma has shown mutant protein accumulation within the endoplasmic reticulum of TM cells, accompanied by increased markers of cellular stress and apoptosis [16]. This degenerative process disrupts the aqueous humor outflow pathway, leading to elevated IOP, and glaucomatous neuropathy. Whole gene deletions and frameshift variants that result in nonsense-mediated decay of the RNA transcript are not associated with glaucoma, as they do not produce abnormal proteins [17]. Pathogenic variants typically involve amino acid substitutions or the generation of truncated proteins [18].

The *MYOC* gene contains three exons encoding a 504-amino acid protein. The Myocilin Database, which compiles glaucoma-related *MYOC* variants, lists 297 coding changes associated with the gene (www.myocilin.com, accessed 25 May 2026) [19]. Most disease-causing variants are located within exon 3, which encodes the olfactomedin domain (amino acid residues 246– 502), a region important for intracellular trafficking [18, 20]. Without cell-based functional evidence such as testing the secretion and/or solubility of variant Myocilin protein in cultured cells, predicting a variant’s pathogenicity is challenging. Variant interpretation involves a thorough review of the literature, summarization of current knowledge, and final variant classification based on these findings. Application of this rigorous process provides diagnostic information that is crucial for determining the best practices for disease management.

Gene variants and their expected pathogenicity can be submitted to the ClinVar database (https://www.ncbi.nlm.nih.gov/clinvar/), which collates interpretations from various sources and highlights discrepancies. Resolving conflicts in variant classification is vital for providing accurate genetic diagnostic information to clinicians, patients, and their families. Standardized guidelines for sequence variant interpretation were published in 2015 by the American College of Medical Genetics and Genomics (ACMG) and the Association for Molecular Pathology (AMP) [21]. Although these recommendations are widely accepted, discrepancies have been reported regarding their application by different laboratories [22, 23, 24]. This has led to the need for disease- and gene-specific guidance from experts in those distinct fields. Within the Clinical Genome Resource (ClinGen, www.clinicalgenome.org)[25], the collaborative Glaucoma Variant Curation Expert Panel (VCEP) was established to review the literature and publicly available evidence on glaucoma and to develop *MYOC*-specific variant interpretation guidelines. In 2022, the Glaucoma VCEP published these specifications, adapting 15 rules to the curation guidelines for *MYOC* and determining that 13 rules were not applicable (version 1.1, 19 November 2021)) [26]. These modified guidelines were piloted on 81 variants, with functional evidence available for 63 (78%), which resulted in classification changes for 40% (16/40) of those listed in ClinVar. A total of 265 variants were then curated and submitted to ClinVar using these guidelines. Here we described the updated *MYOC* specifications developed by the Glaucoma VCEP in 2025, based on more recent literature and revisions from the ClinGen Variant Classification Working Group. We have extended the curation to a total of 271 *MYOC* variants reported in individuals with JOAG/POAG using these latest rules (version 2.1, 6 November 2025, available at https://cspec.genome.network/cspec/ui/svi/doc/GN019). These improvements in variant classification have led to a decrease in the number of variants of uncertain significance (VUS).

## MATERIALS AND METHODS

### Organizational structure of the Glaucoma VCEP

As a part of the ClinGen Ocular Clinical Domain Working Group, the Glaucoma VCEP is responsible for defining specific variant curation rules in accordance with ACMG/AMP guidelines for genes associated with primary glaucoma [27]. The Glaucoma VCEP comprises experts in various fields related to glaucoma, including clinical ophthalmology, research, molecular biology, diagnostics, and genetic counseling. Panel members represent several countries, including the United States, Australia, Germany, and India. Details about the membership can be found on the Glaucoma VCEP website (https://clinicalgenome.org/affiliation/50053/).

### Glaucoma VCEP rule specification and curation process

The first version of the *MYOC* specifications released in November 2021 (v1.1) were used to curate 265 variants [26]. Variants were classified using the ACMG-AMP 5-tier system into Pathogenic (P), Likely Pathogenic (LP), Variant of Uncertain Significance (VUS), Likely Benign (LB) or Benign (B). Initial variant curation assessments were conducted by two biocurators (P.G. and J.H.) using the ClinGen Variant Curation Interface (https://curation.clinicalgenome.org/) to document the applicable rules [28]. Provisional curations were first reviewed by one core approval member (E.S.) and then by two additional core approval members (S.W.T., F.P., K.N.W., K.P.B., or A.D.). A summary of variant classifications was then sent to all Glaucoma VCEP members for feedback before final curation approval. When necessary, the biocurators contacted ClinVar submitters or authors of publications for clarifications on variant nomenclature, affection status, segregation data, or any other lacking information. All variant classifications, applied criteria, and supporting evidence were submitted to ClinVar with a 3 star FDA-level Expert Panel Review status. Variants were annotated using GenBank reference sequences NM_000261.2 for mRNA and NP_000252.1 for protein.

More recently, the specifications were updated (version 2.0 released in December 2024 and version 2.1 in November 2025, available at https://cspec.genome.network/cspec/ui/svi/doc/GN019). Modifications were based on recent publications on variant classifications and updates from the ClinGen Variant Classification Working Group. They were presented and discussed at monthly Glaucoma VCEP online meetings for consensus decisions. These included three updates and four clarifications as described below (with all v2.1 rules detailed in Supplementary Table 1):

Updates:

- PP3/PS1/PM5: Adoption of points capping for the combination of PP3 and PS1 (6 points max) and PP3 and PM5 (5 points max).
- PP3/BP4: Application of REVEL scores at different levels of strength (Supporting, Moderate, Strong) based on calibrated thresholds [31].
- BP4/BP7: Removal of the use of CADD for BP4 and GERP scores for BP7, and application of BP7 to intronic/noncoding/synonymous variants if BP4 is met.

Clarifications:

- Update of MONDO disease ontology term for POAG.
- PS4/PP1: Clarification that individuals with multiple VUS/LP/P variants in *MYOC* are not to be considered.
- PM4: Application to stop-loss variants.
- PM5: Addition of Strong level in the context of 2 previously established P variants.

### Data sources and variant curation

Variants in *MYOC* were selected for curation if they were reported against a glaucoma phenotype in ClinVar, published in a JOAG/POAG case in a scientific journal with DNA level nomenclature, or listed in the Myocilin database (www.myocilin.com)[19]. JOAG was defined as open-angle glaucoma diagnosed before the age of 40. Publicly available variant data were obtained from ClinVar (https://www.ncbi.nlm.nih.gov/clinvar/) between June and November 2021, before the publication of the v1.1 rules in November 2021. Additional published and unpublished case level data were used from research databases of Glaucoma VCEP members with Ethics Committees/IRBs approval, including J.E.C. (The Australian and New Zealand Registry of Advanced Glaucoma [ANZRAG], Southern Adelaide Clinical Human Research Ethics Committee (305.08)) [32], D.A.M. (The Glaucoma Inheritance Study of Tasmania [GIST], Human Research Ethics Committee of Royal Victorian Eye and Ear Hospital (94/237H/19)) [33], F.P. (Ethical review board of the Medical Faculty of the University of Erlangen-Nuremberg (291_12B)), and T.L.Y (Health Sciences Institutional Review Board at the University of Wisconsin-Madison, Wisconsin (2014-1009)). All 265 variants meeting the criteria reported prior to 2022 were curated with v1.1 specifications. Following the release of the v2.1 rules in November 2025, all previously curated variants plus 6 newly reported variants have been reclassified using the v2.1 specifications. Re-curation of variants was performed using the most recent version of gnomAD (v.4.1.0, https://gnomad.broadinstitute.org/). All curated variants are publicly listed in ClinVar with Expert Panel review status and published in the ClinGen Evidence Repository (https://erepo.clinicalgenome.org/evrepo/).

## RESULTS

Version 1.1 of the Glaucoma VCEP rule specifications were applied to the classification of 265 *MYOC* variants, including 81 used in the initial pilot study [26]. This dataset included 245 coding variants published in cases in the literature (including 244 reported in the Myocilin Database and one variant published in a discontinued journal) with associated nucleotide nomenclature and phenotype descriptions reviewed by the Glaucoma VCEP. An additional 20 variants were reported directly in ClinVar by laboratories and were classified against a glaucoma phenotype. A prior ClinVar classification was reported for 27% of the variants (72/265), of which 65% (47/72) maintained overall concordance (B/LB vs VUS vs P/LP) following expert curation (Figure 1). Clinically relevant P/LP classifications had previously been recorded for 19 variants, of which 58% (11/19) retained their status. However, 42% (8/19) of P/LP variants were downgraded; 7 to VUS and 1 to B/LB. In contrast, all 15 variants previously classified as B/LB retained their status. Expert curation reclassified 34% (11/32) of VUS variants to B/LB, but none to P/LP. Conflicting classifications were identified in ClinVar for 8% (6/72) of variants, which expert curation resolved to 5 B/LB and 1 VUS classifications.

**Figure 1:**
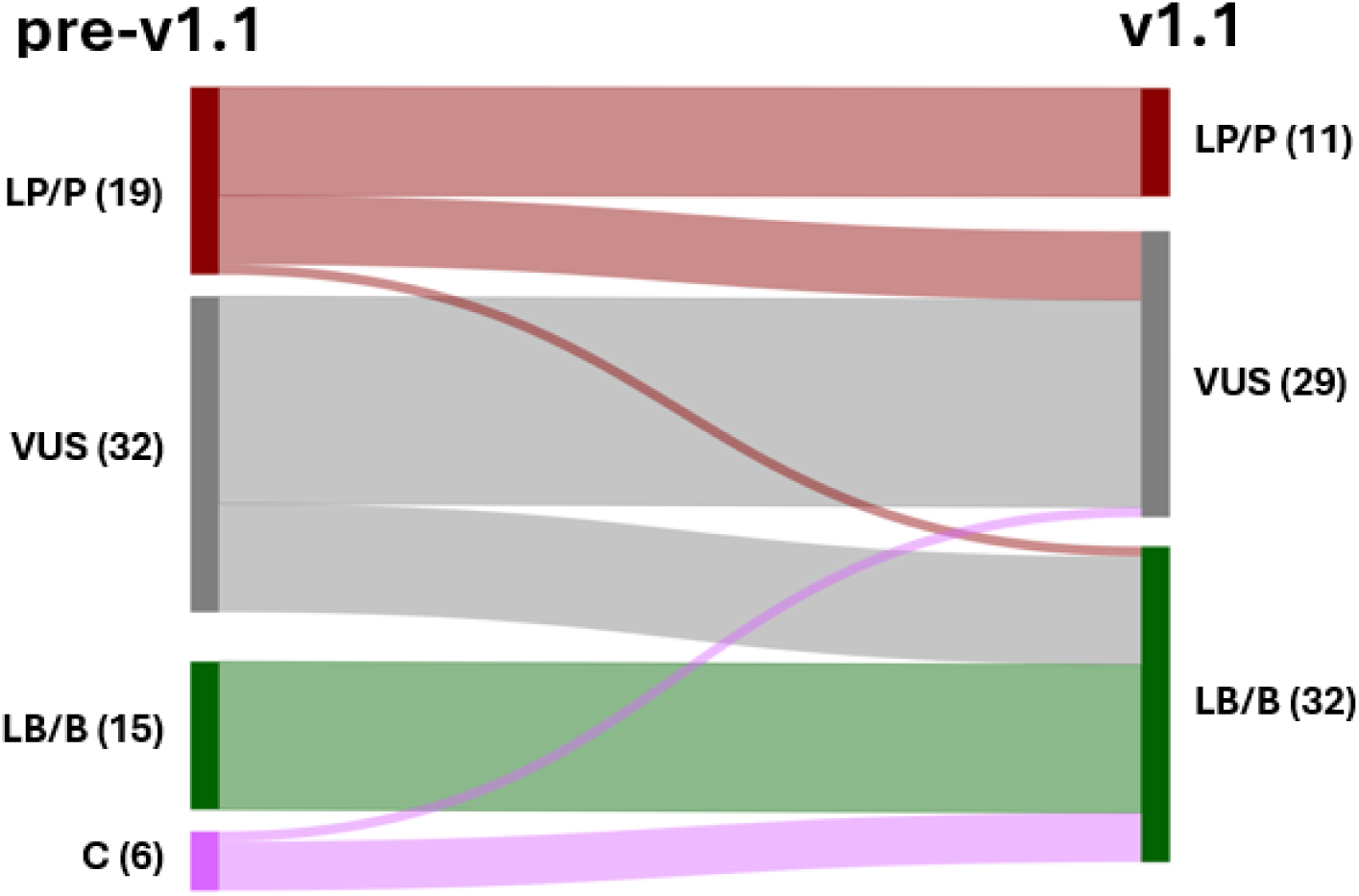
Reclassification of *MYOC* variants within ClinVar using the initial v1.1 rules. Sankey diagram representing the reclassification of variants between no rule specification (pre-v1.1, left) and v1.1 specifications (right). P = pathogenic, LP = likely pathogenic, VUS = variant of uncertain significance, LB = likely benign, B = benign, C = conflicting classifications

The updated version 2.1 *MYOC* specifications were then used to curate the original 265 variants plus an additional 6 variants identified from ClinVar and literature sources. A comparison of variant classifications using v1.1 and v2.1 of the rule specifications is shown in Figure 2. A higher proportion of variants were classified as LP/P and LB/B using v2.1 (105/271=39% vs 74/265=28%). Consequently, there were less VUS classifications using v2.1, with a reduction from 72% to 61%. A total of 43 variants changed classification using the most recent rules, with 95% (41/43) moving to a more clinically relevant B/LB or P/LP classification.

**Figure 2:**
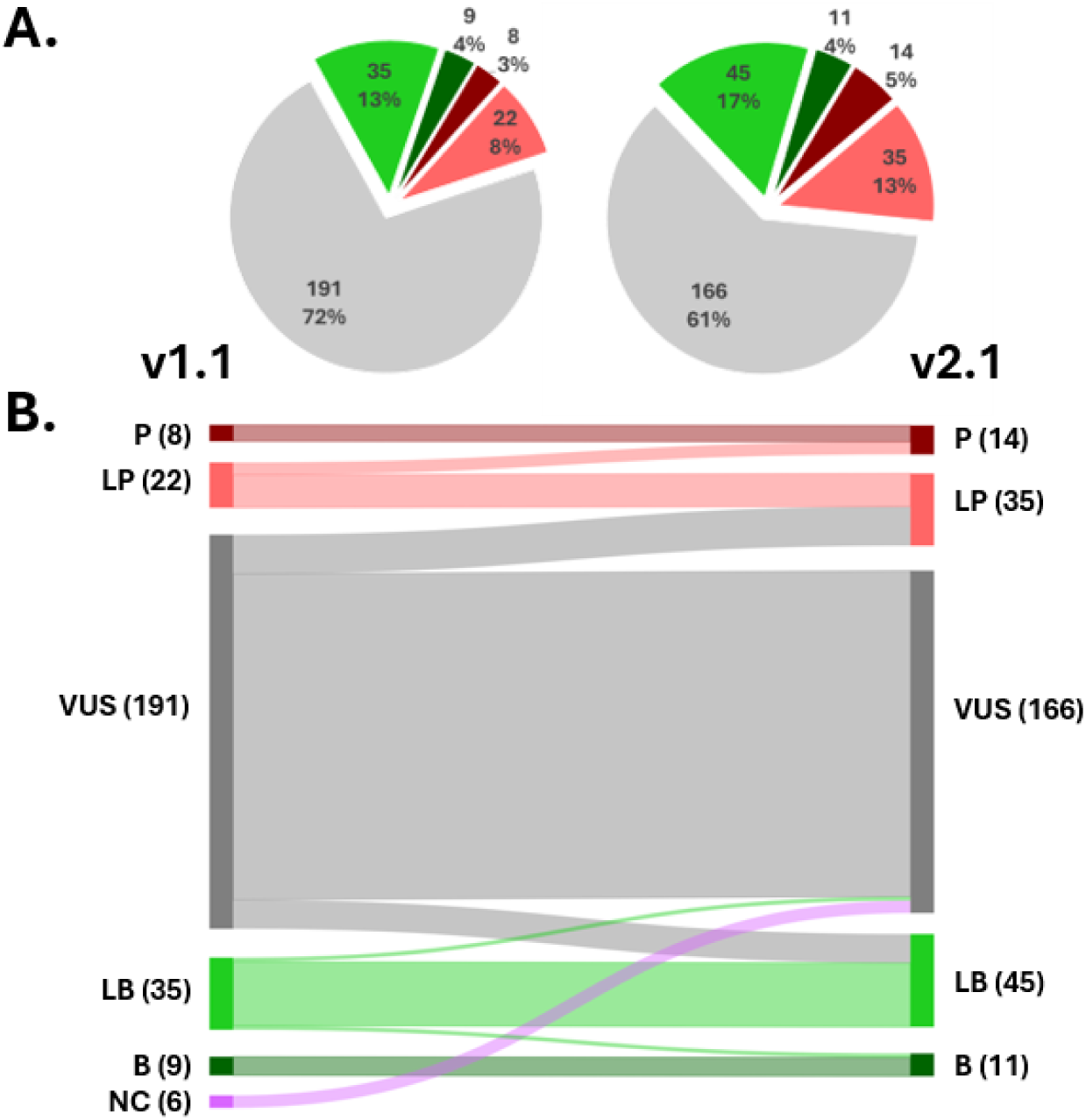
Reclassification of variants using updated *MYOC* rule specifications. A) Pie charts illustrating the proportion of variants assigned to each classification type using specification v1.1 (left) and v2.1 (right). B) Sankey diagram representing the reclassification of variants between the two specification versions. P = pathogenic (dark red), LP = likely pathogenic (red), VUS = variant of uncertain significance (grey), LB = likely benign (light green), B = benign (dark green), NC = new curations.

The distribution of variant types along with their classifications using v2.1 specifications is shown in Figure 3. A full list of the individual variant details is provided in Supplementary Table 2. The majority of variants (68%) were missense, and of these 25% (46/184) were classified as LP/P. Similar numbers of LB/B variants were classified in the missense and synonymous variants groups (25 and 30 variants, respectively), although these variants comprised a higher proportion of the synonymous group (56% of synonymous vs 14% of missense variants). LP/P variants were all located within exon 3, which encodes the conserved olfactomedin domain (Figure 4), and 94% were missense. In contrast, LB/B variants were distributed throughout the entire length of the gene.

**Figure 3:**
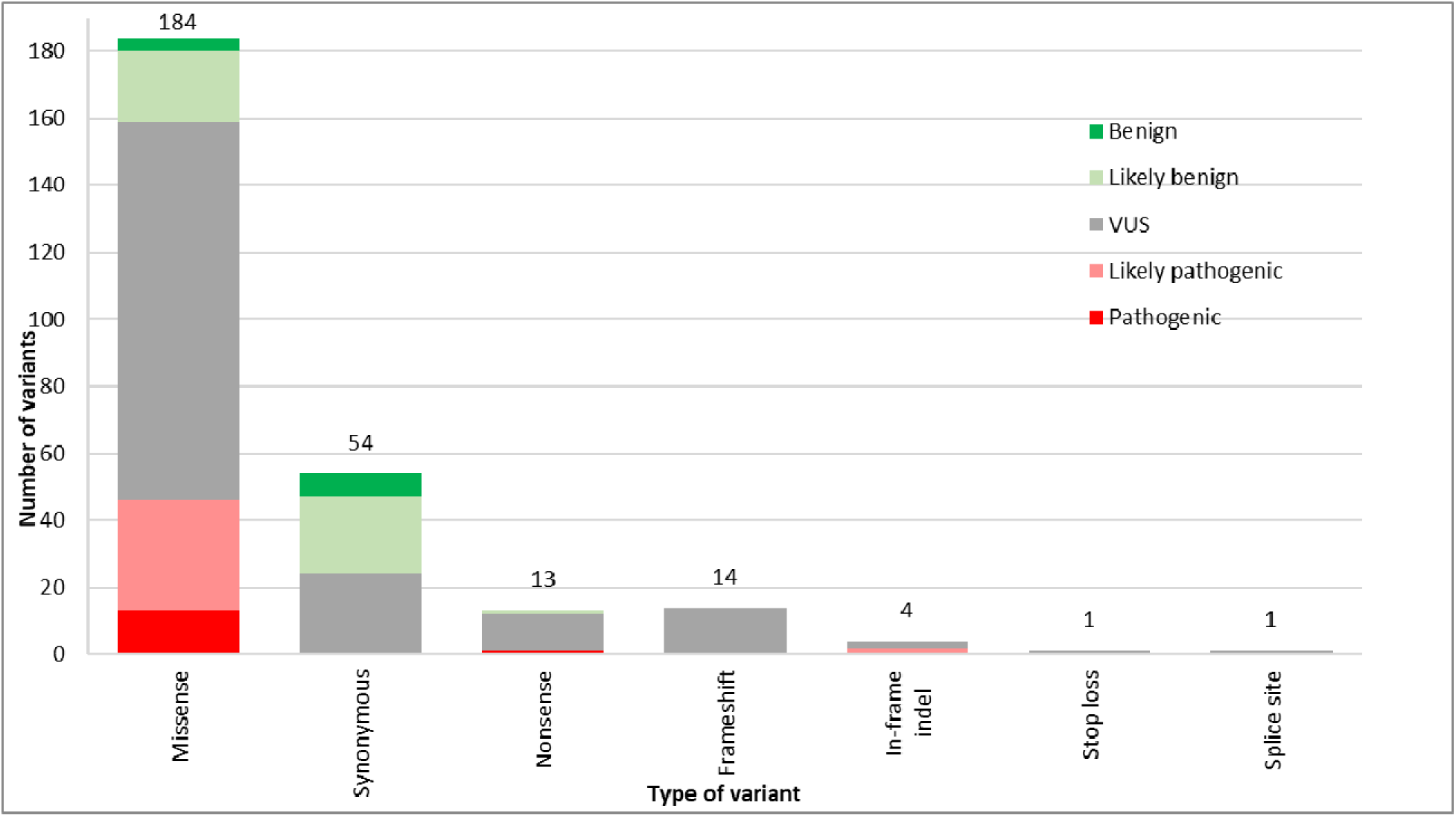
Distribution of classifications by variant type. Columns show the total numbers for each type of variant and their classifications.

**Figure 4:**
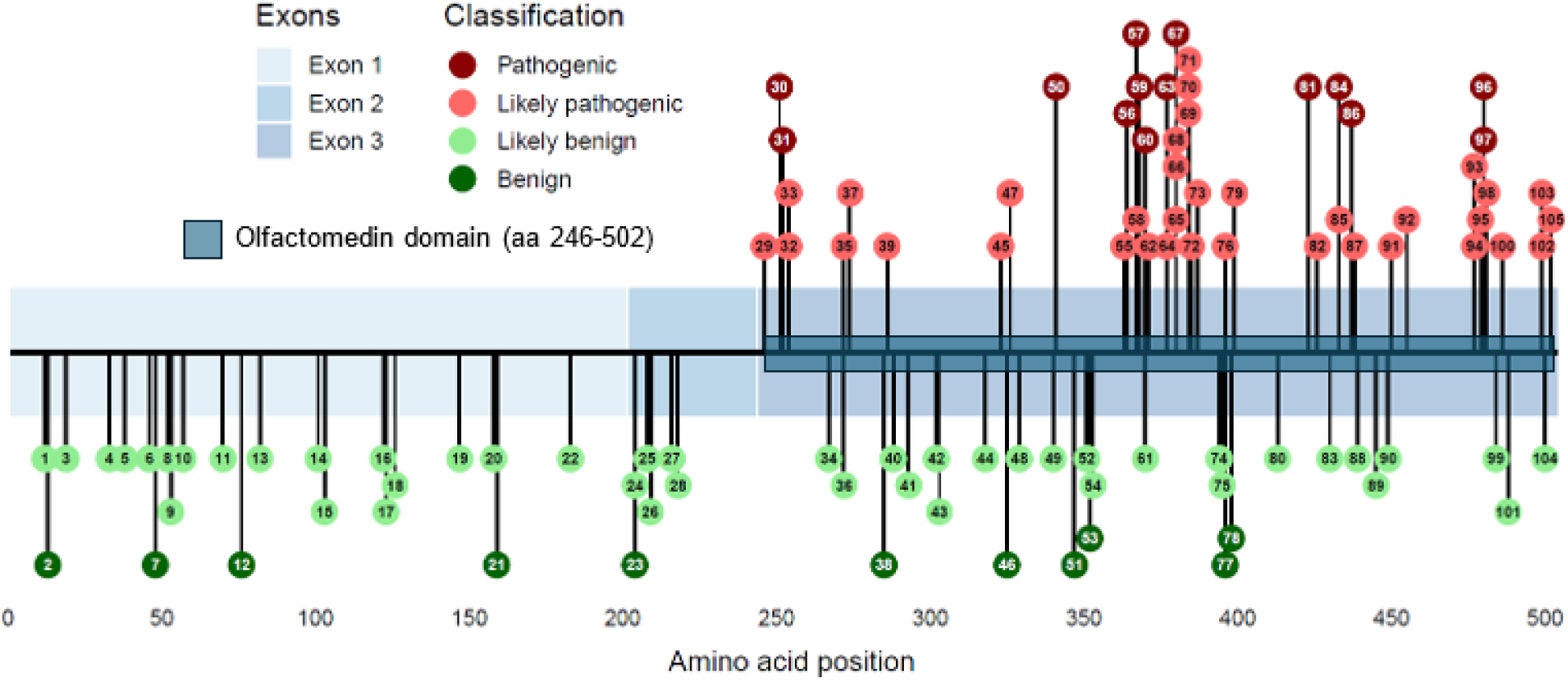
Final classification of non-VUS variants and their genomic location within the *MYOC* gene. Variants classified using v2.1 rules. Exons 1-3 of the *MYOC* gene are represented by blue boxes. Variant gene locations are represented as pins with identifier numbers, as provided in Supplementary Table 2, and classifications color-coded according to the inset key.

The most influential update made by the VCEP from v1.1 to v2.1 was the application of different PP3/BP4 levels of strength based on thresholds of REVEL as the bioinformatic predictor, and the application of BP7 to noncoding and synonymous variants if BP4 is met. With v2.1, the computational criteria (PP3, BP4, BP7) were applied for the first time to 58 variants and at a stronger level to 91 variants (Figure 5, Table 1). These updates led to the reclassification of 10% (19/191) of VUS to LP and 7% (14/191) of VUS to LB.

**Figure 5:**
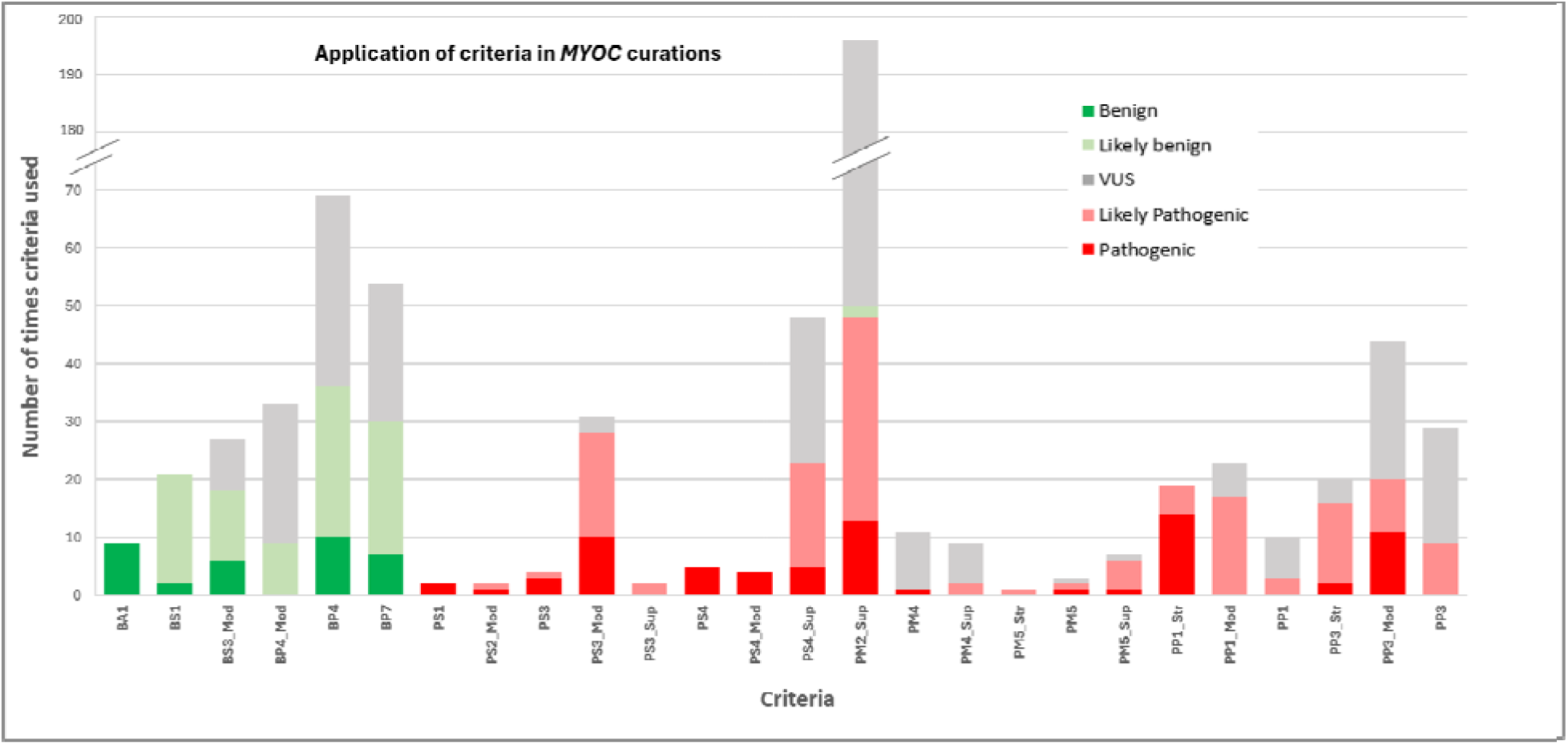
Criteria codes applied during final Glaucoma VCEP classification of variants.

**Table 1:** Impact of computational criteria on the reclassification of MYOC variants. The level at which each computational criterion for PP3, BP4 or BP7 was applied under each version of the rules is shown with the number of variants that changed classification. Notes: BP4 and BP7 may have applied to the same variants. 2 variants reclassified from LB to VUS based on population data were excluded from the table.

|  |  |  | Change in classification |  |  |  |
| --- | --- | --- | --- | --- | --- | --- |
|  |  |  | LB to B | VUS to LB | VUS to LP | LP to P |
| Criteria | v1.1 | v2.1 | n=2 | n=14 | n=19 | n=6 |
| PP3 | Not applied | Not applied |  |  |  |  |
|  |  | PP3 |  |  | 2 |  |
|  | PP3 | PP3 |  |  | 1 |  |
|  |  | PP3_Mod |  |  | 5 | 6 |
|  |  | PP3_Str |  |  | 11 |  |
| BP4 | Not applied | Not applied |  | 1 |  |  |
|  |  | BP4 | 2 | 4 |  |  |
|  |  | BP4_Mod |  | 2 |  |  |
|  | BP4 | BP4 |  | 4 |  |  |
|  |  | BP4_Mod |  | 3 |  |  |
| BP7 | Not applied | Not applied | 2 | 7 |  |  |
|  |  | BP7 |  | 7 |  |  |
|  | BP7 | BP7 |  |  |  |  |

For 9 of the 11 variants classified as B, the stand-alone BA1 criterion (high frequency in population databases) was applied (Figure 5). Additional benign criteria used for these variants included BP4 (computational evidence, 10/11), BP7 (synonymous with no predicted splice site impact, 7/11), BS3_Moderate (functional study evidence, 6/11), and BS1 (allele frequency lower than expected for the disease, 2/9) (Figure 5).

When applying the *MYOC*-specific Glaucoma VCEP rules, the most frequently used criteria was PM2_Supporting (rarity in population databases). This criterion was applied in 72% (196/271) of the classifications, including 13 P, 35 LP, 146 VUS and 2 LB (Figure 5).

Very Strong level criteria are not utilized in the *MYOC*-specified rules. However, 14 variants were classified as P through the combined application of other criteria. All met PP1_Strong (co-segregation with disease), 13/14 met PP3_Strong or Moderate (computational evidence), 13/14 met PS3 or PS3_Moderate (functional evidence) and 9/14 met PS4_Strong or Moderate (prevalence in affected individuals) (Figure 5).

The PS1 criterion (same protein change as an established P variant) was applied only once to the c.1440C>G (p.Asn480Lys) variant, which facilitated its classification as LP (Figure 5). PS2 (*de novo* variant) was applied only twice, to LP/P variants c.752T>C (p.Val251Ala) and c.761C>T (p.Pro254Leu) (Figure 5). PM5 was applied to 11 variants, including at a Supporting level for 7 variants, Moderate level for 3 variants and Strong level for 1 variant (Figure 5).

As part of the curation process, all functional data measuring the disease mechanism that supported the pathogenic or benign nature of *Myocilin* variants were assessed. This included data from animal models which replicated a glaucoma-like phenotype [34, 35, 36, 37], and cell-based protein-overexpression studies that measured Myocilin protein secretion [14, 38, 39, 40, 41], solubility [10, 42], or both [43, 44, 45], and thermal stability [46, 47, 48, 49, 50, 51, 52]. PS3 was applied at a strong level to 4 variants (p.Ser341Pro, p.Pro370Leu, p.Tyr437His, and p.Asn450Tyr) supported by transgenic mouse models [34, 35, 36, 37]. The application of PS3/BS3 was also supported by evidence of non-secretion of variant Myocilin and the accumulation of insoluble aggregates within cells, compared to wild-type protein. The strength of PS3/BS3 application to these assays was based on the odds of pathogenicity (OddsPath) as described by Brnich *et al*. [30]. This required the inclusion of control cells expressing no Myocilin (negative control), wild-type Myocilin (positive control), validated LP/P and LB/B variants, multiple biological and technical replicates, and statistical analyses. None of the assays surpassed an OddsPath score threshold of 18.7 needed for PS3 to be applied at a strong level. Overall PS3_Moderate was applied to 31 variants and BS3_Moderate to 27 variants with sufficient validation controls, while PS3_Supporting was applied to 2 variants with limited evidence supporting protein non-secretion or insolubility.

In instances where functional assays generated conflicting results, recommendations from Brnich *et al*. were used to make decisions [30]. In such cases, the most valid assay overrode the conflicting result, that is the assay with the strongest OddsPath. For instance, if an assay meeting PS3_Moderate showed non-secretion while another meeting PS3_Supporting showed secretion, PS3_Moderate was applied, and vice versa for BS3. When assays with the same level of PS3 or BS3 showed conflicting results, the validity of each assay was reviewed. This approach was used for two variants published by Nakahara et al., where Myocilin over-expressed in human embryonic kidney (HEK-293A) cells was assayed [45]. For p.Asp380His, a Western blot that showed non-secretion and insolubility was considered more reliable than the luciferase assay that showed secretion, leading to the application of PS3_Moderate. For p.Pro481Ser, both assays indicated secretion; however, the Western blot also revealed protein insolubility. Due to these conflicting results, PS3 or BS3 was not applied. Additionally, when the most valid assay showed only partial solubility or secretion, PS3 or BS3 was not applied.

Functional evidence was available for 33% (90/271) of all variants, but it could only be applied to 24% (64/271). VCEP interpretation guidelines excluded functional evidence where there were insufficient validation controls (variants reaching a LB/B or LP/P classification without functional evidence) to meet the OddsPath threshold, or where there were inconsistent results or methodology that prevented combining assays from the same class. For the 64 variants with functional data applied, final classifications were 13 P, 21 LP, 12 VUS, 12 LB and 6 B, resulting in 81% (52/64) with a clinically relevant classification. This clearly contrasts with the overall 39% (105/271) of variants with clinically relevant classifications, underscoring the need for additional functional evidence to reclassify current VUS.

Out of all VUS, 93% (154/166) lacked functional evidence or were associated with assay results that could not be applied. Among these, 43% (66/154) had a total score of 0 or -1 points and could be reclassified as LB if BS3 were applied (Figure 6). Similarly, 12% (19/154) of these VUS had scores of 4 or 5 points, and could be reclassified as LP if PS3 were applied (Figure 6). Overall, additional functional data could reclassify 51% (85/166) of VUS as more clinically relevant LB or LP variants.

**Figure 6:**
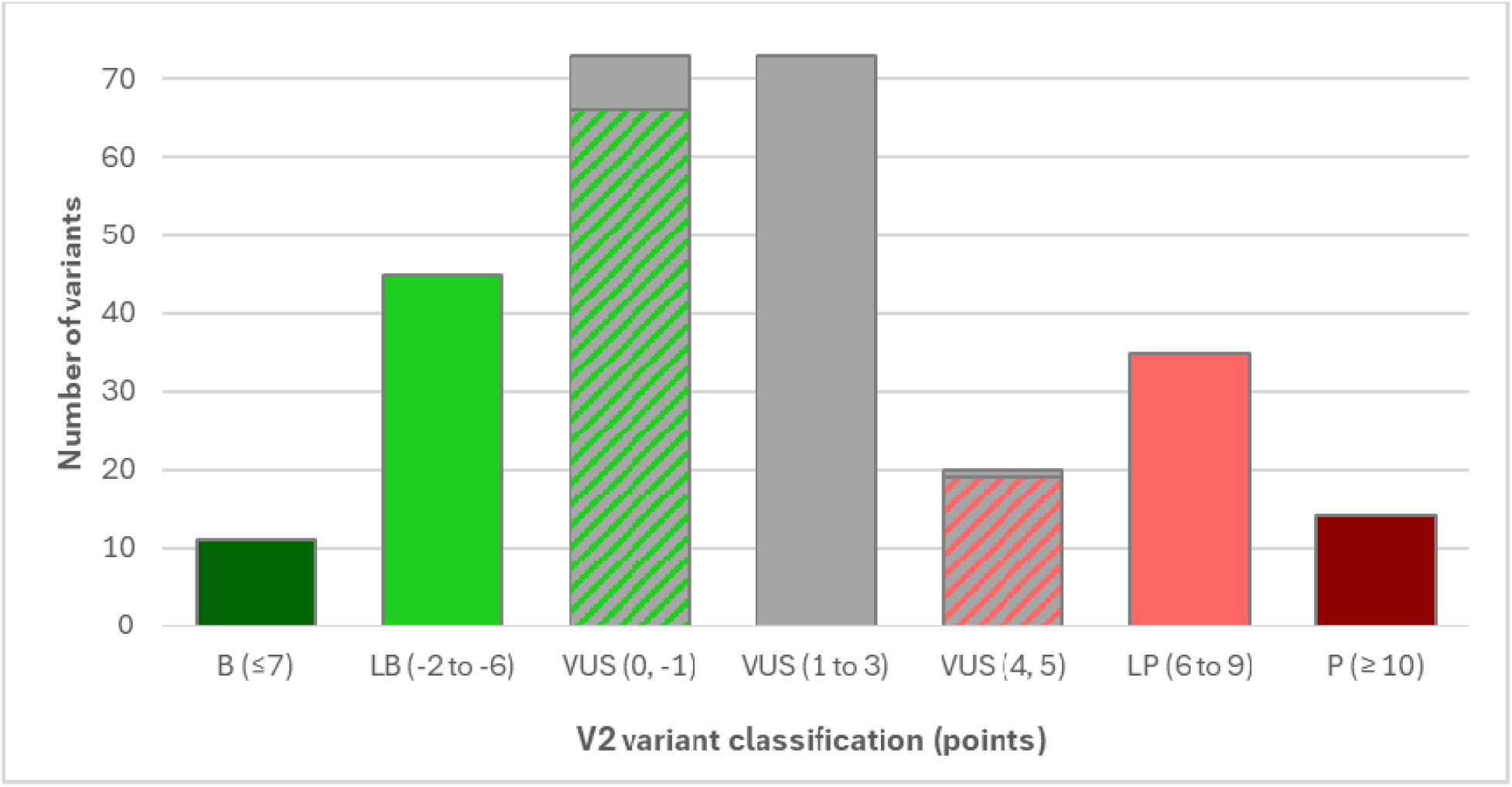
Current classification and potential impact of additional functional evidence on VUS. Striped columns represent variants which could potentially change classification to LB (green) or LP (red).

Despite these advancements, 72% (138/193) of variants submitted to the ClinVar database by the Glaucoma VCEP or others since November 2021 were classified as VUS. This reflects a general lack of evidence supporting pathogenicity for most variants reported in the scientific literature or by testing laboratories. Significantly, 19% (31/166) of VUS have solely been reported by testing laboratories in ClinVar for individuals that are unaffected, have an unknown affection status, or have been associated with a phenotype unrelated to the *MYOC* gene. This highlights the strong need for public database submissions to include associated phenotypic data that will improve our ability to classify variants.

## DISCUSSION

In this study, we evaluated the impact of iterative refinements to *MYOC* specific ACMG/AMP specifications developed by the Glaucoma VCEP through application to a large and diverse set of *MYOC* variants. Implementation of the updated v2.1 *MYOC* specifications resulted in a notable shift toward more clinically definitive classifications, with 39% of variants classified as LP/P or LB/B compared with 28% under v1.1. Importantly, 95% of reclassified variants moved toward greater clinical relevance, underscoring that these changes did not introduce classification inflation but instead resolved previous uncertainty (reduced proportion of VUS). This improvement largely reflects refinement of computational evidence thresholds and the structured application of BP7 to synonymous and noncoding variants, supporting the value of quantitative bioinformatic calibration. These findings demonstrate that updates to disease- and gene specific interpretation guidelines can meaningfully impact the clinical relevance of variant classifications within the ACMG/AMP framework and highlight areas where additional evidence generation will have the greatest influence for *MYOC* variants.

Consistent with the established disease mechanism of *MYOC* associated glaucoma, all LP/P variants were confined to exon 3, encoding the olfactomedin domain. This domain-specific enrichment reinforces the central role of olfactomedin misfolding and impaired secretion in *MYOC* related disease pathogenesis. In contrast, LB/B variants were distributed throughout the gene, reflecting both mutational tolerance outside the olfactomedin domain and the contribution of synonymous and population frequent variants to benign classifications. These observations further validate the biological plausibility of the resulting classifications.

Adjustments to computational evidence application represented the most impactful change between specification versions. Rather than expanding reliance on *in silico* predictions, v2.1 introduced calibrated strength thresholds for PP3 and BP4 based on REVEL performance and the systematic application of BP7, restricting their application to contexts with biological plausibility. These computational criteria were newly applied or strengthened for a large fraction of variants and directly contributed to the reclassification of nearly one fifth of VUS to LP or LB. This approach aligned with ClinGen recommendations for cautious integration of computational evidence and demonstrates how quantitative thresholds can reduce uncertainty without disproportionately weighting prediction tools and overestimating pathogenicity.

Functional evidence proved to be a critical determinant of clinically definitive classifications. While functional data were available for 33% of variants, only rigorously validated assays meeting standardized evidence weighting were applied, resulting in functional evidence contributing to classification for 24% of variants. By restricting use of PS3 and BS3 to assays meeting OddsPath based thresholds, many published functional datasets were deliberately excluded, prioritizing reliability over completeness. This highlights an ongoing challenge in variant interpretation: the need for community wide adoption of standardized functional validation frameworks. Among variants with applied functional data, 81% reached a clinically relevant classification, compared with only 39% across the full dataset. This striking contrast underscores the disproportionate value of well validated functional assays in resolving *MYOC* VUS.

The stringent application of PS3/BS3 criteria, including the exclusion of assays lacking adequate controls or exhibiting conflicting results, reflects a deliberate balance between evidence utilization and avoidance of misclassification. Conflicting functional results were addressed using established OddsPath based prioritization, ensuring that higher quality evidence superseded less reliable assays. This approach proved essential in cases where secretion and solubility assays yielded discordant results and highlights the importance of standardized validation requirements. Without such rigor, conflicting functional data risks introducing, rather than resolving, uncertainty in variant interpretation.

The persistence of a high proportion of VUS reflects structural limitations in the available evidence rather than deficiencies in the interpretation framework itself. In particular, the lack of validated functional assays and incomplete phenotypic annotation for rare variants continue to constrain classification across genes and disease areas. Numerous VUS demonstrated evidence that approached, but did not meet, thresholds for LB or LP classification. Our analyses suggest that over half of current VUS could be resolved with appropriately validated functional data, emphasizing that evidence scarcity, not rule inadequacy, remains the principal barrier to definitive classification. Furthermore, many unresolved VUS were derived from ClinVar submissions lacking detailed phenotypic context, limiting the application of segregation and case control criteria. Improved standards for expanded phenotype reporting alongside variant submissions would substantially enhance interpretability of rare *MYOC* variants and accelerate downstream resolution of uncertain classification efforts across ClinGen resources. These findings mirror challenges reported by other VCEPs and underscore the evidence limited nature of contemporary variant catalogs.

Collectively, these findings have important clinical implications. Enhanced *MYOC*-specific specifications improve confidence in variant interpretation, facilitating appropriate genetic counseling and risk assessment for individuals undergoing glaucoma testing. From a research perspective, our results prioritize the development of standardized, high throughput functional assays targeting Myocilin secretion, solubility, and stability, which would have immediate impact on VUS resolution.

In conclusion, this study demonstrates how systematic refinement of gene specific ACMG/AMP specifications can measurably improve variant classification outcomes while maintaining conservative evidentiary standards and methodological rigor. Although uncertainty remains for many variants, our findings clearly demonstrate that targeted functional evidence generation and improved phenotype reporting represent the most effective strategies for further reducing the burden of VUS in *MYOC* associated glaucoma. Beyond *MYOC*, these findings provide a practical example of how iterative rule calibration, rigorous functional evidence vetting, and transparent curation practices can advance consistency and clinical utility of variant interpretation efforts across genes and disorders.

## Data Availability

All variant curations are published in ClinVar and in the ClinGen Evidence Repository.

https://erepo.genome.network/evrepo/ui/summary/classifications?columns=gene&values=MYOC&matchTypes=contains&pgSize=25&pg=1&matchMode=or

https://www.ncbi.nlm.nih.gov/clinvar/search/?gene=myoc&assembly=GRCh38

## ACKNOWLEDGMENTS

This study was supported by a National Health and Medical Research Council (NHMRC) of Australia Centre for Program Grant [GNT1150144], Practitioner Fellowships to J.E.C. [GNT2026787], D.A.M. and A.W.H., and Investigator grant to E.S. [GNT2016545], a Snow Medical Research Foundation to O.M.S. [PF2019-040], a Core Grant for Vision Research from the National Eye Institute (NEI) of the National Institutes of Health (NIH) to the University of Wisconsin Madison [P30EY016665] and an Unrestricted Grant from Research to Prevent Blindness, Inc. to the UW Madison Department of Ophthalmology and Visual Sciences to S.W.T., K.N.W. and T.L.Y., the Peter A. Duehr Professorship to T.L.Y., and a NEI/NIH grant [R21EY034251] to S.W.T. ClinGen is primarily funded by the National Human Genome Research Institute (NHGRI) with co-funding from the National Cancer Institute (NCI), through the following grants: Baylor/Stanford [U24HG009649], Broad/Geisinger [U24HG006834], and UNC/Kaiser [U24HG009650]. The content is solely the responsibility of the authors and does not necessarily represent the official views of the National Institutes of Health.

## WEB RESOURCES

CADD score (v1.6): https://cadd.gs.washington.edu/snv

ClinGen: https://clinicalgenome.org/

ClinGen Evidence Repository: (https://erepo.clinicalgenome.org/evrepo/

ClinGen Glaucoma VCEP: https://www.clinicalgenome.org/affiliation/50053/

ClinGen Variant Curation Interface: https://curation.clinicalgenome.org/

ClinVar: https://www.ncbi.nlm.nih.gov/clinvar/

GERP score: https://genome.ucsc.edu/cgi-bin/hgGateway

gnomAD: https://gnomad.broadinstitute.org/

Myocilin database: http://www.myocilin.com

REVEL score: https://sites.google.com/site/revelgenomics/downloads

SpliceAI score: https://spliceailookup.broadinstitute.org/

Whiffin/Ware calculator: https://cardiodb.org/allelefrequencyapp/

## SUPPORTING INFORMATION

Additional supporting information can be found online in the Supporting Information section at the end of this article.

All variant classifications presented in this article have been submitted to ClinVar by the Glaucoma VCEP. Refer to Supplementary Table 2 for a list of all variants and their associated ClinVar IDs.

**Supplementary Table 1**: Summary of updated Glaucoma VCEP rule specifications and pathogenicity points applicable to *MYOC* variants. Adapted from Burdon et al., 2022 [26]. Strength options and modifications are shown for each criterion, e.g., PP1 can be applied at different strength levels, i.e., Supporting, Moderate or Strong, whereas PM2 is only applied at the Supporting level (PM2_Supporting) and not at a Moderate level as the criterion name suggests. *BA1 is a stand-alone criterion to meet a Benign variant classification. ^#^The combination of PP3 and PM5 should not be higher than 5 points, and the combination of PP3 and PS1 should not be higher than 6 points. Updated rules between v1.1 and v2.1 are shown in bold. Points are assigned based on each of the applied criterion strengths and summated to arrive at a final classification, as proposed by Tavtigian et al., 2020 [29]. JOAG = Juvenile open-angle glaucoma, POAG = Primary open-angle glaucoma

**Supplementary Table 2: Details of 271 *MYOC* variants classified by the Glaucoma VCEP using v2.1 rule specifications.** Variants, ClinVar IDs, applicable specification criteria, total scores, and their final classification are provided. Points are assigned based on each of the applied criterion strengths and summated to arrive at a final classification, as proposed by Tavtigian et al., 2020 [29]. SA = Stand Alone. N/A = Not applicable, BA1 is a standalone criterion for Benign, therefore points do not apply to variants meeting BA1.

