## Supplementary Table 1 for "ClinGen Glaucoma Variant Curation Expert Panel recommendations enhance classification of myocilin variants"

| **Criterion** | **Strength Options** | **Points** | **Category** | **Specifications in v2.1** |
| --- | --- | --- | --- | --- |
| PS1 | PS1  PS1_Moderate | 4  2 | Known P/LP missense change resulting from different DNA change. | Same amino acid change as established P (Strong) or LP (Moderate). Must be assessed independently of PS1, must not affect splicing (SpliceAI < 0.2).^#^ |
| PS2 | PS2_Very strong  PS2  PS2_Moderate  PS2_Supporting | 8  4  2  1 | *De novo* in an affected individual and no family history. | ≥ 2 confirmed de novo in JOAG (Strong), ≥ 2 confirmed de novo in POAG or 1 confirmed de novo in JOAG or ≥2 assumed de novo in JOAG (Moderate), 1 confirmed de novo in POAG or ≥ 2 assumed de novo in  POAG or 1 assumed de novo in JOAG (Supporting). Both parents must have no clinical signs of glaucoma. |
| PS3 | PS3  PS3_Moderate  PS3_Supporting | 4  2  1 | Well-established *in vitro* or *in vivo* functional studies supportive of a damaging effect. | Thermostability, solubility or secretion assays or animal models replicating glaucoma phenotype. Assays with OddsPath > 18.7 (Strong), OddsPath > 4.3 (Moderate), or OddsPath > 2.1 (Supporting; see Brnich 2019 [30]). |
| PS4 | PS4  PS4_Moderate  PS4_Supporting | 4  2  1 | Prevalence of variant in affected individuals is significantly increased over controls. | ≥ 15 (Strong), ≥ 6 (Moderate), or ≥ 2 (Supporting) probands from multiple studies clinically diagnosed with JOAG/POAG. PM2_Supporting must be met. |
| PM2 | PM2_Supporting | 1 | Allele frequency is ≤ 0.01% in population databases. | Highest allele frequency in any general population database of ≥ 10,000 alleles. |
| PM4 | PM4  PM4_Supporting | 2  1 | Change to protein length. | In-frame indels, **stop-loss** and truncating variants involving ≥ 10% (Moderate) or < 10% (Supporting) of the protein, located within the olfactomedin domain (residues 246-502). |
| PM5 | **PM5_Strong**  PM5  PM5_Supporting | 4  2  1 | Different missense change at the same codon as known pathogenic missense variant. | Missense variant at the same residue as 2 established P variants (Strong), 1 P or 2 LP variants (Moderate) or 1 LP variant (Supporting). Must be assessed independently of PM5, must not affect splicing (SpliceAI ≤ 0.2), meets PP3 and has a Grantham score ≥ established P/LP variants.^#^ |
| PP1 | PP1_Strong  PP1_Moderate  PP1 | 4  2  1 | Co-segregation with disease in multiple affected family members. | Co-segregating meioses ≥ 7 in > 1 family (Strong), ≥ 5 (Moderate), and ≥ 3 (Supporting). Affected individuals must be clinically assessed and have relevant signs or confirmed glaucoma. Obligate carriers can be counted. BA1 and BS1 must not be met. |
| PP3 | **PP3_Strong**  **PP3_Moderate**  PP3 | 4  2  1 | Computational evidence supports a deleterious effect. | **Missense variants with REVEL score ≥ 0.932 (Strong), 0.773-0.931 (Moderate) or 0.644-0.772 (Supporting).^#^** |
| BA1 | BA1 | n/a* | Allele frequency is ≥ 1% in population databases. | Highest allele frequency in any general population databases of ≥ 2,000 alleles and be present in ≥ 5 alleles. |
| BS1 | BS1 | -4 | Allele frequency is ≥ 0.1% in population databases. | As for BA1. Does not apply to p.Gln368Ter. |
| BS3 | BS3_Moderate  BS3_Supporting | -2  -1 | Well-established *in vitro* or *in vivo* functional studies show no effect. | Applies to variants assessed for thermostability, solubility or secretion in functional assays for studies with OddsPath < 0.23 (Moderate) or < 0.48 (Supporting; see Brnich 2019 [30]). |
| BP4 | **BP4_Strong**  **BP4_Moderate**  BP4 | -4  -2  -1 | Computational evidence supports no impact. | **Missense variants with REVEL score ≤ 0.016 (Strong), 0.017-0.183 (Moderate) or 0.184-0.290 (Supporting). Synonymous, non-coding or intronic variants with SpliceAI score ≤ 0.1.** |
| BP7 | BP7 | -1 | Synonymous variant for which no predicted impact on splicing. | **Intronic, noncoding or synonymous exonic variant if BP4 is met.** |
